# Bringing COVID-19 home for Christmas: a need for enhanced testing in healthcare institutions after the holidays

**DOI:** 10.1101/2020.12.18.20248460

**Authors:** David R. M. Smith, Audrey Duval, Jean Ralph Zahar, Lulla Opatowski, Laura Temime, on behalf of the EMEA-MESuRS working group on nosocomial SARS-CoV-2 modelling

**Affiliations:** Institut Pasteur, Epidemiology and Modelling of Antibiotic Evasion (EMAE), Paris, France; Université Paris-Saclay, UVSQ, Inserm, CESP, Anti-infective evasion and pharmacoepidemiology team, Montigny-Le-Bretonneux, France; Modélisation, épidémiologie et surveillance des risques sanitaires (MESuRS), Conservatoire national des arts et métiers, Paris, France; IAME, UMR 1137, Université Paris 13, Sorbonne Paris Cité, France; Service de Microbiologie Clinique et Unité de Contrôle et de Prévention du Risque Infectieux, Groupe Hospitalier Paris Seine Saint-Denis, AP-HP, Bobigny, France; PACRI unit, Institut Pasteur, Conservatoire national des arts et métiers, Paris, France

## Abstract

Festive gatherings this 2020 holiday season threaten to cause a surge in new cases of novel coronavirus disease 2019 (COVID-19). Hospitals and long-term care facilities are key hotspots for COVID-19 outbreaks, and may be at elevated risk as patients and staff return from holiday celebrations in the community. Some settings and institutions have proposed fortified post-holiday testing regimes to mitigate this risk. We use an existing model to assess whether implementing a single round of post-holiday screening is sufficient to detect and manage holiday-associated spikes in COVID-19 introductions to the long-term care setting. We show that while testing early helps to detect cases prior to potential onward transmission, it likely to miss a substantial share of introductions owing to false negative test results, which are more probable early in infection. We propose a two-stage post-holiday testing regime as a means to maximize case detection and mitigate the risk of nosocomial COVID-19 outbreaks into the start of the new year. Whether all patients and staff should be screened, or only community-exposed patients, depends on available testing capacity: the former will be more effective, but also more resource-intensive.

## Main

As much of the world gathers to celebrate Christmas and the end of the 2020 calendar year, there is mounting concern that a holiday-associated surge in novel coronavirus disease 2019 (COVID-19) lies in wait for early 2021. Past experiences in China, Israel and elsewhere have demonstrated a need to prepare for spikes in COVID-19 incidence associated with festive holidays, which tend to draw individuals from distant places into close contact for prolonged periods.^1,2^ Nevertheless, in a compassionate bid to alleviate the pandemic’s psychological toll, healthcare authorities in some regions will relax their COVID-19 restrictions this holiday season. In France, for instance, the Ministry of Health announced that nursing homes will admit visitors and allow residents to return to the community to stay with their families, provided they undergo RT-PCR or antigenic testing upon their return.^3^

For healthcare facilities in areas with active community circulation of SARS-CoV-2, post-holiday screening regimes seem well-founded. Yet surveillance is challenged by several factors, including delayed/absent clinical symptoms and imperfect diagnostic sensitivity of gold-standard RT-PCR tests, which are unlikely to detect SARS-CoV-2 over the first three days of infection.^4^ For long-term care residents and healthcare workers returning to their respective institutions after the holidays, and potentially exposed to the virus all the while, a simple test upon their return may not suffice.

We adapted an existing COVID-19 surveillance model to evaluate post-holiday screening regimes in the long-term care setting. Described elsewhere,^5^ this model uses inter-individual contact data to reproduce patient-staff interactions in a rehabilitation hospital, and simulates transmission and clinical progression of SARS-CoV-2 infection, as well as active RT-PCR testing with time-varying diagnostic sensitivity, peaking at 80% eight days post-infection.^4^ Here, we made minor adjustments to reflect the current epidemiological context: (i) baseline immunizing seroprevalence among 20% of patients and staff, and (ii) reinforced infection control practices (e.g. face masks), reducing per-contact transmission risk by 80%.^6^

We initialized our model to reflect enhanced community SARS-CoV-2 acquisition risk for 100% of staff and 50% of patients over the holidays (Figure 1A). This translated to three members of staff and one patient returning to the hospital with “holiday-associated” SARS-CoV-2 infection at simulation outset, with an on average 5-day age of infection (range 1-8). An additional 0-3 holiday-associated cases were gradually introduced from the community over the following week. Ensuing nosocomial outbreaks were simulated for two weeks (Figure 1B). We evaluated a suite of mass-testing strategies deployed at varying time intervals over the week following Christmas, under the baseline assumption that individuals presenting with COVID-19-like symptoms are tested immediately (Figure 1C). For each strategy, we calculated the proportion of imported cases detected and the cumulative number of secondary cases averted over two weeks, assuming that infected individuals stop transmitting once diagnosed, and accounting for a 24-hour delay for RT-PCR test results (Figure 1B).

**Figure 1.**
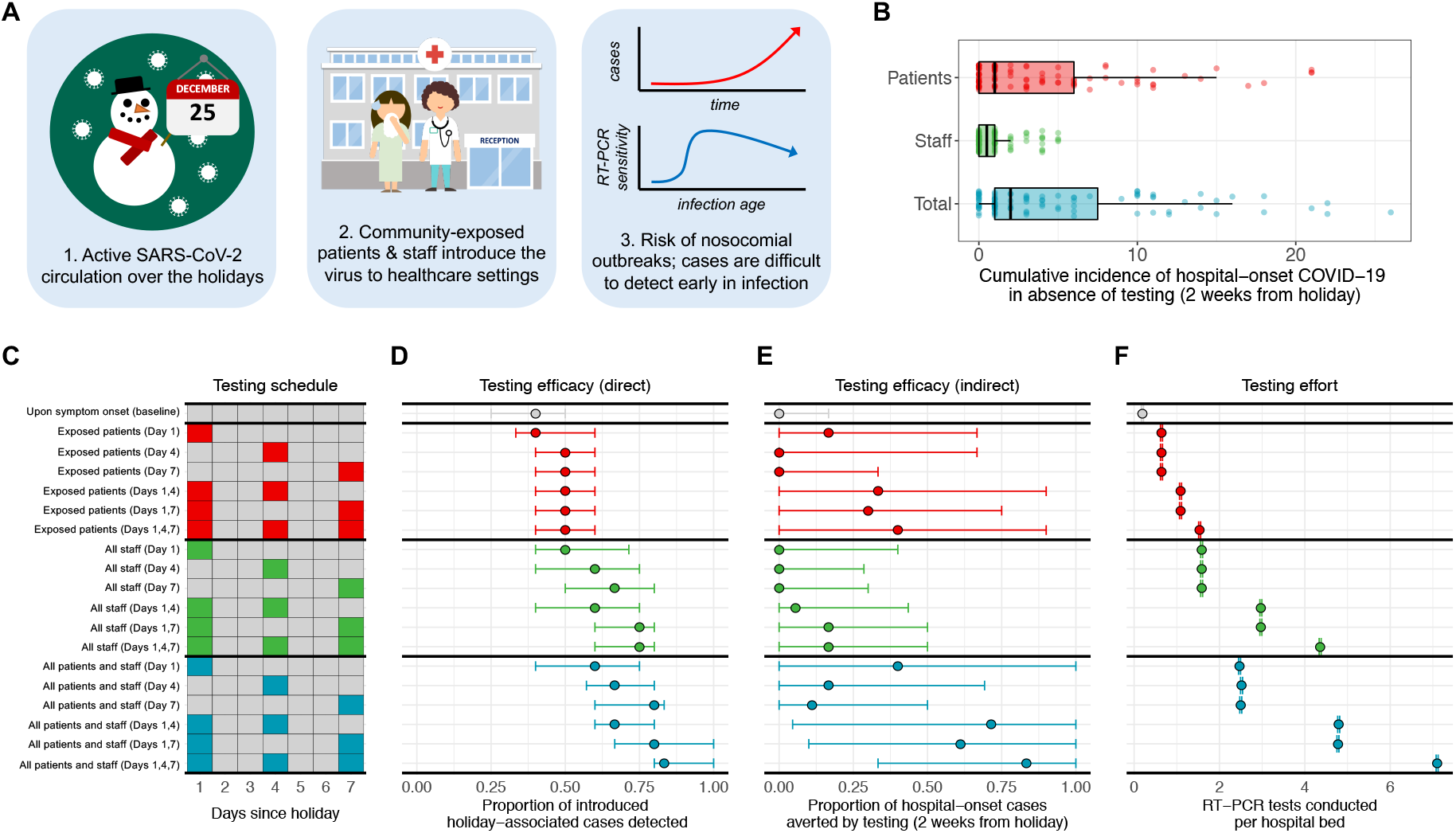
Mass-testing regimes can prevent COVID-19 outbreaks triggered by a surge in holiday-associated community incidence. (A) Festive celebrations among patients and staff can result in introductions of holiday-associated COVID -19 cases to healthcare institutions, potentially triggering nosocomial outbreaks. Detecting new cases is challenged by time-varying sensitivity of RT-PCR. (B) Simulated cumulative incidence of COVID-19 among patients and staff within two weeks of first introduction of holiday-associated cases, among a hospital population averaging 240 staff and 170 patients per week. (C) Proposed schedules for RT-PCR testing, measured as days since return from holiday. Coloured strategies represent mass-screening events conducted in addition to an assumed baseline strategy of testing all patients and staff upon presentation of COVID-19-like symptoms. (D) Proportion of imported holiday-associated cases detected by each testing strategy within the first week from holiday. (E) Proportion of downstream nosocomial cases averted by testing strategies, measured up to two weeks from holiday and excluding simulations in which no nosocomial transmission occurred. (F) Number of RT-PCR tests conducted under each strategy. For D-F, dots represent medians and bars represent the interquartile range computed over 10,000 stochastic simulations.

We found distinct advantages to different mass-testing schedules. When conducted earlier (1 day since return from holiday), testing was more likely to detect imported cases prior to transmission events, thereby preventing a greater share of onward nosocomial spread (Figure 1E). However, imported cases were more likely to be missed, because not all introductions were necessarily present on day 1, and because false-negative tests were overall more likely at earlier time points given the time-varying nature of RT-PCR sensitivity (Figure 1D). Taken together, multi-day screening regimes were most effective for both detecting holiday-associated SARS-CoV-2 introductions and preventing onward nosocomial spread.

Screening everyone present in the hospital was unsurprisingly more effective than screening only patients or staff, but also more resource-intensive (Figure 1F). Given resource limitations, testing community-exposed patients was found to avert more cases than testing only staff. This result may be biased by the characteristics of the hospital we modelled, which includes more staff than patients and high rates of patient-to-patient contact.^7^ Rapid antigen testing may help to alleviate resource limitations, but is less diagnostically sensitive than RT-PCR, and perhaps more likely to benefit from multi-stage screening to overcome false-negative results.

After a fraught 2020, many will find solace in reuniting with their loved ones this holiday season. This may come at the unfortunate cost of energizing the spread of SARS-CoV-2, with potential to trigger outbreaks in hospitals, nursing homes and other vulnerable settings. Promising vaccines on the horizon may turn the tide in 2021, but are unlikely to have substantial epidemiological impact before the new year. In the meantime, with COVID-19 cases at greater numbers worldwide than ever before, healthcare institutions should consider enhanced post-holiday surveillance regimes to protect their patients and staff from infection.

## Data Availability

Simulated data is available from authors upon reasonable request.

## Acknowledgments

The members of the EMEA-MESuRS working group on nosocomial SARS-CoV-2 modeling are Audrey Duval, Kévin Jean, Sofía Jijón, Ajmal Oodally, Lulla Opatowski, George Shirreff, David RM Smith and Laura Temime.

